# Social Determinants of Health and Preventive Service Utilization in a U.S. Community Health Center: An Application of the Andersen and Aday Model

**DOI:** 10.64898/2026.01.14.26344123

**Authors:** Jennifer M. Torres Del Valle, Miguel Ángel Sánchez González, Marisol Peña Orellana, José Carrión Baralt

**Affiliations:** School of Medicine, Complutense University of Madrid, Madrid, Spain; Graduate School of Public Health, Medical Sciences Campus, University of Puerto Rico, San Juan, Puerto Rico

## Abstract

**Objective:** To examine how social determinants of health (SDOH) influence the utilization of preventive health services at a Federally Qualified Health Center (FQHC) in Erie, Pennsylvania, using the Andersen and Aday behavioral model (Andersen, 1995; Aday & Andersen, 1974).

**Methods:** A cross-sectional, descriptive correlational study was conducted among adults (≥18 years) receiving primary care at Community Health Net from March through October 2018. The data was collected via a self-administered questionnaire, classifying variables as predisposing, enabling, or need factors, per the Andersen and Aday model. The Protocol for Responding to & Assessing Patients’ Assets, Risks & Experiences (PRAPARE) tool was used to assess SDOH. Mental health status was measured using the Patient Health Questionnaire-9 and Generalized Anxiety Disorder 7-item scale. Logistic regression identified predictors of preventive service utilization.

**Results:** Of the 274 participants, 65.8% were over 50 years old, 50.2% had low annual incomes (≤$15,000), 71.0% had public insurance, 58.8% were unemployed, and 10.0% were homeless. A total of 78.8% of the participants reported experiencing emotional or physical problems that affected their social life; 16.1% reported having transportation barriers and 13.9% reported a lack of social support. Preventive service utilization was high (75.1%). Multivariable analysis showed that low incomes (≤$15,000) (odds ratio [OR] = 1.82, 95% CI [1.15, 2.89]), unemployment (OR = 2.10, 95% CI [1.30, 3.40]), and mental health conditions (OR = 3.05, 95% CI [1.80, 5.18]) were significant predictors of utilization. Homelessness showed a positive but not statistically significant association (OR = 1.68, 95% CI [0.92, 3.06). The most frequently reported unmet needs among participants were mental health care (24.8%), transportation (16.1%), and social support (13.9%).

**Conclusions:** Socioeconomic disadvantage and mental health needs are strongly associated with preventive service utilization in this FQHC population, consistent with the Andersen and Aday model and international evidence (Braveman & Gottlieb, 2014; Marmot et al., 2008; Magnan, 2021). Addressing SDOH—including poverty, unemployment, mental health, and access barriers— remains essential for advancing health equity in high-risk communities (World Health Organization, 2008; Artiga & Hinton, 2022).

## Introduction

Health is profoundly shaped by the conditions in which people are born, grow, live, work, and age; these conditions are collectively known as SDOH (World Health Organization, 2008; Marmot et al., 2008). Decades of research have established that factors such as poverty, education, employment, housing, and access to transportation are as important as perhaps more important than—clinical care in determining health outcomes and life expectancy (Braveman & Gottlieb, 2014; Magnan, 2021; Solar & Irwin, 2010).

In the United States, persistent health disparities are closely linked to social and structural inequities, disproportionately affecting low-income, racial and ethnic minority, and other marginalized populations (Williams et al., 2019; Artiga & Hinton, 2022). For example, individuals living in poverty have higher rates of chronic disease, lower preventive service utilization, and shorter lifespans compared to their more advantaged counterparts (Woolf et al., 2015; Daniel et al., 2018). FQHCs play a critical role in addressing these inequities by providing comprehensive, community-based care to underserved populations (Shi et al., 2012). However, even within FQHCs, barriers such as the lack of transportation, unstable housing, and limited social support can impede access to preventive and primary care (De Marchis et al., 2019; Centers for Disease Control and Prevention, 2023).

The Andersen and Aday behavioral model remains one of the most widely used frameworks for understanding healthcare utilization, emphasizing the interplay of predisposing, enabling, and need factors (Andersen, 1995; Aday & Andersen, 1974). Recent research and policy initiatives— including the Healthy People 2030 framework and the National Academy of Medicine’s “Five Plus Five” approach—underscore the urgent need to systematically address SDOH in all healthcare settings (Office of Disease Prevention and Health Promotion, 2020; Magnan, 2021).

Despite growing recognition of the multiple effects of SDOH, there remains a critical need to build an evidence base through empirical studies that quantify said effects in real-world, high-risk populations and inform actionable strategies for health systems and policymakers (Braveman et al., 2022; Artiga & Hinton, 2022). This study contributes to that evidence base by using validated tools and a robust conceptual model to identify associations between multiple SDOH and preventive service utilization in a U.S. FQHC.

### Methodology

A cross-sectional, descriptive correlational study was conducted from March through October 2018 at Community Health Net (CHN), an FQHC in Erie, Pennsylvania. A total of 274 adults aged 18 years or older who received primary care at CHN were recruited through consecutive sampling. The inclusion criteria were 18 years old or older, receiving primary care at CHN, and signing an informed consent.

Data were collected using a self-administered questionnaire, and study variables were categorized according to the Andersen and Aday behavioral model of health service utilization:

- **Predisposing factors:** age, gender, race/ethnicity, education attainment.
- **Enabling factors:** insurance status, employment status, income, housing stability, social support, transportation access.
- **Need factors:** presence of chronic conditions and mental health status.

SDOH were assessed using the Protocol for Responding to and Assessing Patients’ Assets, Risks, and Experiences (PRAPARE) tool (National Association of Community Health Centers, 2023). Mental health status was evaluated using the Patient Health Questionnaire-9 (PHQ-9) to assess depressive symptoms and the Generalized Anxiety Disorder 7-item scale (GAD-7) to assess anxiety (De Marchis et al., 2019).

Univariate, bivariate, and multivariable analyses were conducted, and logistic regression models were used to identify predictors of preventive service utilization. All analyses were performed using SAS statistical software version 9.4. Statistical significance was defined **as** p < 0.05.

## Results

The sample was characterized by high rates of poverty, unemployment, public insurance coverage, and housing instability. Additionally, 16.1% of the participants reported having transportation barriers and 13.9% reported a lack of social support. Emotional and/or physical problems affecting the participant’s social life were also prevalent, as reported by 78.8% of the sample. Of the 274 participants, 206 (75.1%) reported at least one primary care visit in the previous year. Table 1 presents the sociodemographic characteristics of the study sample.

**Table 1.**
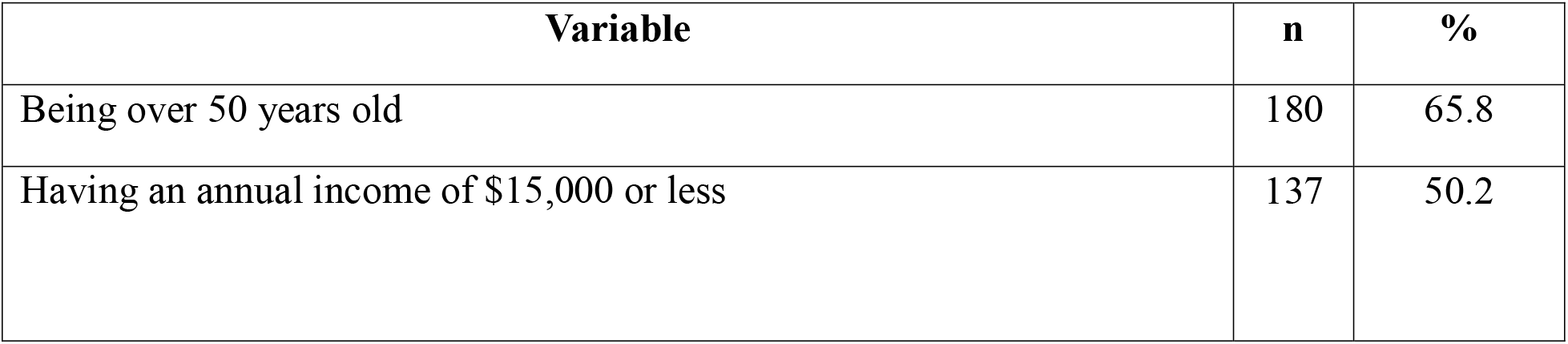
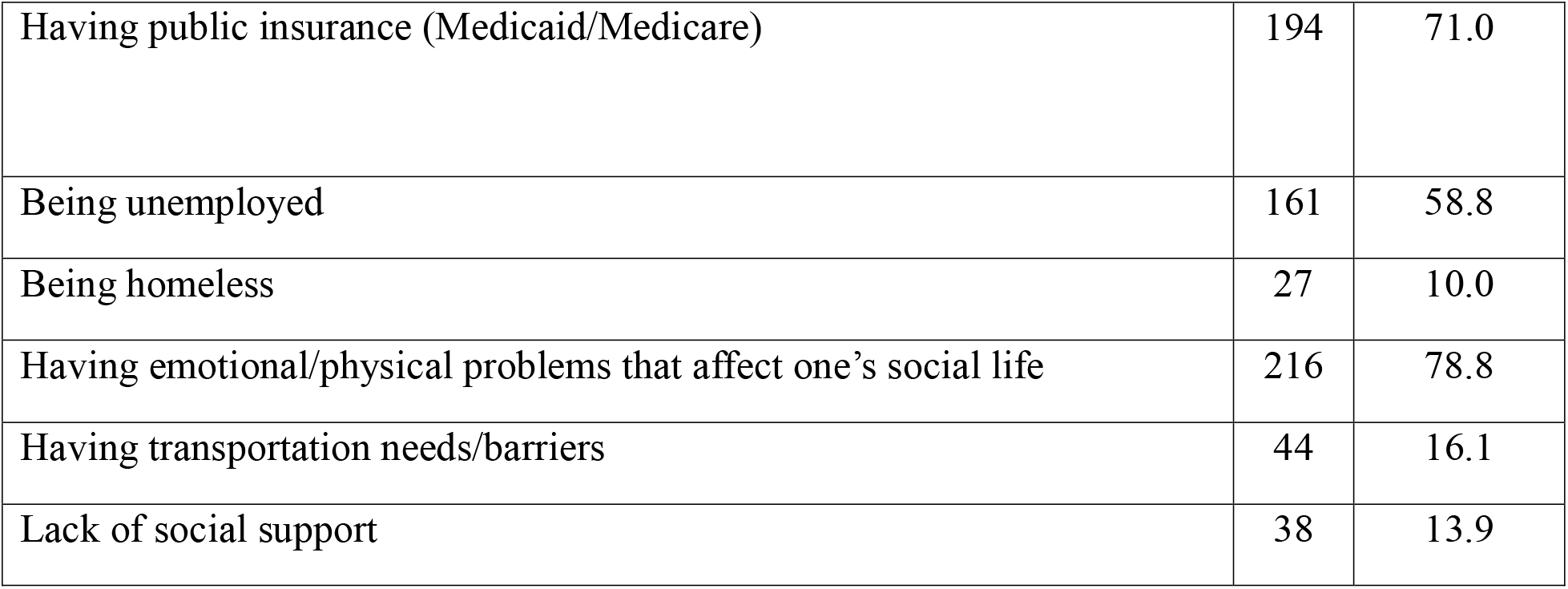
Sociodemographic characteristics of study participants.

### Predictors of Preventive Service Utilization

Multivariable logistic regression analysis identified several social determinants as significant predictors of preventive service utilization:

- **Income** ≤ **$15**,**000** was associated with higher odds of using preventive services (OR = 1.82, 95% CI [1.15, 2.89], *p* = .011).
- **Unemployment** was also a strong predictor (OR = 2.10, 95% CI [1.30, 3.40], *p*= .002).
- **Presence of a mental health condition** was associated with over three times the odds of utilization (OR = 3.05, 95% CI [1.80, 5.18], *p* < .001).
- **Homelessness** showed a positive though not statistically significant association (OR = 1.68, 95% CI [0.92, 3.06], *p* = .091).

Multivariable analysis further demonstrated that the presence of a mental health condition was the strongest predictor of preventive service utilization, with an odds ratio of 3.05 (95% CI: 1.80–5.18, *p* < .001). Results from the multivariable logistic regression are shown in Table 2.

**Table 2.** Multivariable logistic regression results for predictors of preventive service utilization.

| Predictor | Odds Ratio | 95% CI | P value |
| --- | --- | --- | --- |
| Having an income of \$15,000 or less | 1.82 | 1.15–2.89 | .011 |
| Being unemployed | 2.10 | 1.30–3.40 | .002 |
| Having a mental health condition | 3.05 | 1.80–5.18 | <.001 |
| Being homeless | 1.68 | 0.92–3.06 | .091 |

These findings are consistent with those of previous research demonstrating that socioeconomic disadvantage and mental health needs are associated with increased healthcare utilization, particularly in safety-net settings (Braveman & Gottlieb, 2014; Marmot et al., 2008; Magnan, 2021).

### Predictors of Unmet Needs

As shown in Table 3, multivariable logistic regression analysis identified several social determinants of health associated with higher odds of reporting unmet needs. The presence of a mental health condition was associated with the greatest odds of unmet needs (OR = 3.05, 95% CI: 1.80–5.18, *p* < .001). Lower income (≤ $15,000) was also significantly associated with unmet needs (OR = 1.82, 95% CI: 1.15–2.89, *p* = .011), as was unemployment (OR = 2.10, 95% CI: 1.30–3.40, *p* = .002). Enrollment in Medicaid or Medicare was associated with higher odds of unmet needs (OR = 1.55, 95% CI: 1.03–2.34, *p* = .037). Homelessness showed a positive but not statistically significant association with unmet needs (OR = 1.68, 95% CI: 0.92–3.06, *p* = .091), and age over 50 years was not significantly associated with unmet needs (OR = 1.22, 95% CI: 0.80–1.87, *p* = .350).

**Table 3.** Odds ratios for predictors related to unmet needs and social determinants.

| Predictor | Odds Ratio | 95% CI | P value |
| --- | --- | --- | --- |
| Having a mental health condition | 3.05 | 1.80–5.18 | <.001 |
| Having an income of \$15,000 or less | 1.82 | 1.15–2.89 | .011 |
| Being unemployed | 2.10 | 1.30–3.40 | .002 |
| Being enrolled in Medicaid/Medicare | 1.55 | 1.03–2.34 | .037 |
| Being homeless | 1.68 | 0.92–3.06 | .091 |
| Being over 50 years old | 1.22 | 0.80–1.87 | .350 |

## Discussion

This study quantifies the impact of SDOH on preventive service utilization in a vulnerable population. Notably, individuals with low annual incomes (≤$15,000) had 1.8 times the odds of using preventive services compared with those with higher incomes, with unemployment doubling the likelihood of utilization. The presence of a mental health condition was the strongest predictor, tripling the odds of accessing preventive care. These findings align with national and international research demonstrating that socioeconomic disadvantage, mental health needs, and social instability are key drivers of health service use and shape health outcomes (Braveman & Gottlieb, 2014; Marmot et al., 2008; Magnan, 2021).

Also, the study suggests that individuals experiencing mental health challenges were more than three times as likely to seek preventive health services than were those without such conditions. These results are consistent with those of the broader literature, which indicates that mental health needs often drive increased healthcare utilization, particularly in low-income and underserved communities where barriers to specialized care are common.

The high prevalence of unmet mental health needs observed in this study aligns with prevalence rates found in national and international research, highlighting the intersection between mental health, SDOH, and healthcare access. Factors such as poverty, unemployment, and the lack of social support not only increase the risk of mental health problems but also exacerbate difficulties in accessing appropriate care (Braveman & Gottlieb, 2014; Marmot et al., 2008). In this context, the absence of integrated mental health services within primary care settings further compounds disparities and limits opportunities for early intervention.

Taken together, addressing mental health as a core component of primary care—through systematic screening, referral pathways, and partnerships with community organizations—is essential for improving health outcomes and reducing inequities in populations served by community health centers. The findings of this study underscore the urgent need to expand access to mental health services as part of a comprehensive strategy to address SDOH and promote equity in healthcare utilization.

While homelessness was associated with higher odds of service use, this association did not reach statistical significance, possibly due to the limited sample size or unmeasured confounders. Transportation barriers and the lack of social support, though prevalent, were not directly measured as predictors in the regression, but their high frequency among unmet needs underscores their relevance to intervention planning (De Marchis et al., 2019).

These results reinforce the Andersen and Aday model’s emphasis on enabling and need factors (Andersen, 1995; Aday & Andersen, 1974) and support recommendations from the World Health Organization and Healthy People 2030 to address SDOH through policy and practice (World Health Organization, 2008; Office of Disease Prevention and Health Promotion, 2020). Interventions targeting SDOH—including transportation assistance, social support programs, and integrated mental health care—have demonstrated effectiveness in reducing disparities and improving outcomes in high-need populations (Braveman et al., 2022; Artiga & Hinton, 2022).

## Conclusions

This study demonstrates that SDOH—particularly poverty, unemployment, mental health conditions, and social support—play a critical role in shaping preventive service utilization among patients at a specific FQHC in the United States. The findings reveal that individuals with lower income, those who are unemployed, and those with mental health conditions are significantly more likely to access preventive services, highlighting the complex interplay between need, vulnerability, and healthcare-seeking behavior (Braveman & Gottlieb, 2014; Marmot et al., 2008).

The high prevalence of unmet needs for mental health care, transportation, and social support underscores persistent barriers to comprehensive care in vulnerable populations. While the presence of public insurance facilitates some access, it does not guarantee adequate quality of life or access to all necessary services, particularly specialized or supportive care. These results reinforce the Andersen and Aday behavioral model’s assertion that enabling and need factors are central to healthcare utilization (Andersen, 1995; Aday & Andersen, 1974).

From a policy perspective, these findings support the urgent need for integrated, multisectoral interventions that address both the clinical and the non-clinical determinants of health. Expanding access to mental health services, improving transportation infrastructure, and strengthening social support networks should be prioritized in FQHCs and similar settings (De Marchis et al., 2019; Artiga & Hinton, 2022). Furthermore, public insurance programs should be reviewed to ensure they cover a sufficiently broad range of services, including those that address SDOH and behavioral health. For clinicians and health-system leaders, routine screening for SDOH using validated tools such as PRAPARE should become standard practice, with clear referral pathways and community partnerships to address identified needs (National Association of Community Health Centers, 2023; Magnan, 2021). These strategies align with national initiatives such as Healthy People 2030 and global recommendations from the World Health Organization)

Finally, future research should continue to explore the mechanisms by which SDOH influence healthcare utilization and outcomes, including the roles of intersectionality, the presence of systemic barriers, and the effectiveness of targeted interventions. Longitudinal studies and implementation research are essential to inform best practices and policy reforms that can close gaps in care and reduce health disparities in the United States and beyond (Braveman et al., 2022; Williams et al., 2019).

In summary, addressing SDOH is not only a moral imperative but also a practical necessity for improving preventive service utilization and advancing health equity in high-risk communities.

## Future Directions

Future research should expand on this study’s findings by exploring how the integration of mental health services within primary care settings influences access to care and service utilization among medically underserved populations, particularly in FQHCs. Given the associations observed between socioeconomic disadvantages such as poverty, unemployment, and public insurance coverage—and unmet mental health needs, future studies should prioritize the development and evaluation of interventions that address social determinants of health in a coordinated manner. Longitudinal research designs will be essential to better understand how social risk factors shape mental health service utilization over time. Additionally, examining the role of community partnerships and social support mechanisms may provide further insight into strategies for reducing barriers to care and improving health-related quality of life in underserved communities.

## Limitations

The cross-sectional design of this study enabled the identification of associations between social determinants of health and preventive service utilization; however, it limits the ability to infer causality. Specifically, the temporal ordering and directionality of the observed associations cannot be established. Future research using longitudinal study designs is needed to better characterize causal pathways and to examine how social risk factors influence patterns of health service utilization over time.

## Ethics Statement

The research protocol was reviewed and approved by the Research Committee of the School of Medicine at the Complutense University of Madrid. All study procedures were conducted in accordance with the ethical standards outlined in the Declaration of Helsinki. Written informed consent was obtained from all participants prior to data collection.

## Data Availability Statement

The data supporting the findings of this study are not publicly available due to ethical and privacy restrictions but are available from the corresponding author upon reasonable request.

## Funding Statement

This work was supported by the Hispanics-In-Research Capability Endowment (HiREC) Program funded by the National Institute on Minority Health and Health Disparities (NIMHD), National Institutes of Health (NIH) (Grant No. S21MD001830), and partially supported by the NIH-funded Clinical and Translational Research Training Program (R25MD007607). The content is solely the responsibility of the authors and does not necessarily represent the official views of the National Institutes of Health. The funders had no role in the study design, data collection, analysis, interpretation, or manuscript preparation.

## Notes

### Competing Interest Statement

The authors have declared no competing interest.

### Author Declarations

The Research Committee of the School of Medicine of the Complutense University of Madrid gave ethical approval for this work.

## References

Aday, L. A., & Andersen, R. M. (1974). A framework for the study of access to medical care. Health services research, 9(3), 208–220. https://www.ncbi.nlm.nih.gov/pmc/articles/PMC1071804/

Andersen, R. M. (1995). Revisiting the behavioral model and access to medical care: Does it matter? Journal of health and social behavior, 36(1), 1–10. 10.2307/2137284

Artiga, S., & Hinton, E. (2022). Beyond health care: The role of social determinants in promoting health and health equity. Kaiser Family Foundation. Published May 10, 2022. https://www.kff.org/racial-equity-and-health-policy/issue-brief/beyond-health-care-the-role-of-social-determinants-in-promoting-health-and-health-equity/

Braveman, P., & Gottlieb, L. (2014). The social determinants of health: It’s time to consider the causes of the causes. Public health reports (Washington, D.C.: 1974), 129 Suppl 2(Suppl 2), 19–31. 10.1177/00333549141291S206

Braveman, P., Arkin, E., Proctor, D., Kauh, T., & Holm, N. (2022). Systemic and structural racism: Definitions, examples, health damages, and approaches to dismantling. Health affairs (Project Hope), 41(2), 171–178. 10.1377/hlthaff.2021.01394

Centers for Disease Control and Prevention. (2023). Social determinants of health: Know what affects health. Published January 17, 2024. https://www.cdc.gov/socialdeterminants/index.htm

Daniel, H., Bornstein, S. S., & Kane, G. C., Health and Public Policy Committee of the American College of Physicians, Carney, J. K., Gantzer, H. E., Henry, T. L., Lenchus, J. D., Li, J. M., McCandless, B. M., Nalitt, B. R., Viswanathan, L., Murphy, C. J., Azah, A. M., & Marks, L. (2018). Addressing social determinants to improve patient care and promote health equity: An American College of Physicians position paper. Annals of internal medicine, 168(8), 577–578. 10.7326/M17-2441

De Marchis EH, Hessler D, Fichtenberg C, Fleegler EW, Huebschmann AG, Clark CR, Cohen AJ, Byhoff E, Ommerborn MJ, Adler N, Gottlieb LM. Assessment of Social Risk Factors and Interest in Receiving Health Care-Based Social Assistance Among Adult Patients and Adult Caregivers of Pediatric Patients. JAMA Netw Open. 2020 Oct 1;3(10):e2021201. doi: 10.1001/jamanetworkopen.2020.21201. PMID: 33064137; PMCID: PMC7568201.

Gottlieb LM, Wing H, Adler NE. A Systematic Review of Interventions on Patients’ Social and Economic Needs. Am J Prev Med. 2017 Nov;53(5):719–729. doi: 10.1016/j.amepre.2017.05.011. Epub 2017 Jul 5. PMID: 28688725.

Magnan, S. (2021). Social determinants of health 101 for health care: Five plus five. [Discussion paper] NAM Perspectives. National Academy of Medicine. 10.31478/202104c

Marmot, M., Friel, S., Bell, R., Houweling, T. A., & Taylor, S. (2008). Closing the gap in a generation: Health equity through action on the social determinants of health. Lancet (London, England), 372(9650), 1661–1669. 10.1016/S0140-6736(08)61690-6

National Association of Community Health Centers. (2023). PRAPARE: Protocol for Responding to and Assessing Patients’ Assets, Risks, and Experiences. https://prapare.org/

Office of Disease Prevention and Health Promotion. (2020). Healthy People 2030: Social determinants of health. https://health.gov/healthypeople/objectives-and-data/social-determinants-health

Shi, L., Lebrun, L. A., & Tsai, J. (2012). Assessing the impact of the health center growth initiative on health center patients. Public health reports (Washington, D.C.: 1974), 127(1), 66–75. 10.1177/003335491012500215

Solar, O., & Irwin, A. (2010). A conceptual framework for action on the social determinants of health. Social Determinants of Health Discussion Paper 2 (Policy and Practice).World Health Organization. https://www.who.int/publications/i/item/9789241500852

Torres Del Valle, J. M. (2024). Determinantes sociales de salud, utilización de servicios preventivos y calidad de vida en un centro de salud comunitaria de los Estados Unidos [Doctoral dissertation, Universidad Complutense de Madrid].

Williams, D. R., Lawrence, J. A., & Davis, B. A. (2019). Racism and health: Evidence and needed research. Annual review of public health, 40, 105–125. 10.1146/annurev-publhealth-040218-043750

Woolf, S. H., Aron, L. Y., Dubay, L., Simon, S. M., Zimmerman, E., & Luk, K. X. (2015). How are income and wealth linked to health and longevity? Urban Institute and Center on Society and Health. https://www.urban.org/research/publication/how-are-income-and-wealth-linked-health-and-longevity

World Health Organization. (2008). Closing the gap in a generation: Health equity through action on the social determinants of health. Final Report of the Commission on Social Determinants of Health. https://www.who.int/publications/i/item/WHO-IER-CSDH-08.1

